# Application of Optical Genome Mapping For Comprehensive Assessment of Chromosomal Structural Variants for Clinical Evaluation of Myelodysplastic Syndromes

**DOI:** 10.1101/2021.01.13.21249611

**Authors:** Hui Yang, Guillermo Garcia-Manero, Diana Rush, Guillermo Montalban-Bravo, Saradhi Mallampati, L. Jeffrey Medeiros, Brynn Levy, Rajyalakshmi Luthra, Rashmi Kanagal-Shamanna

**Affiliations:** Department of Leukemia, The University of Texas MD Anderson Cancer Center, Houston, TX; Department of Hematopathology, The University of Texas MD Anderson Cancer Center, Houston, TX; Department of Pathology, Columbia University Medical Center, New York, NY

**Keywords:** Optical genome mapping, myelodysplastic syndrome, karyotype, microarray, karyotype, Bionano imaging, leukemia

## Abstract

Structural chromosomal variants [copy number variants (CNVs): losses/ gains and structural variants (SVs): inversions, balanced and unbalanced fusions/translocations] are important for diagnosis and risk-stratification of myelodysplastic syndromes (MDS). Optical genome mapping (OGM) is a novel single-platform cytogenomic technique that enables high-throughput, accurate and genome-wide detection of all types of clinically important chromosomal variants (CNVs and SVs) at a high resolution, hence superior to current standard-of-care cytogenetic techniques that include conventional karyotyping, FISH and chromosomal microarrays. In this proof-of-principle study, we evaluated the performance of OGM in a series of 12 previously well-characterized MDS cases using clinical BM samples. OGM successfully facilitated detection and detailed characterization of twenty-six of the 28 clonal chromosomal variants (concordance rate: 93% with conventional karyotyping; 100% with chromosomal microarray). These included copy number gains/losses, inversions, inter and intra-chromosomal translocations, dicentric and complex derivative chromosomes; the degree of complexity in latter aberrations was not apparent using standard technologies. The 2 missed aberrations were from a single patient within a composite karyotype, below the limit of detection. Further, OGM uncovered 6 additional clinically relevant sub-microscopic aberrations in 4 (33%) patients that were cryptic by standard-of-care technologies, all of which were subsequently confirmed by alternate platforms. OGM permitted precise gene-level mapping of clinically informative genes such as *TP53, TET2* and *KMT2A*, voiding the need for multiple confirmatory assays. OGM is a potent single-platform assay for high-throughput and accurate identification of clinically important chromosomal variants.

## INTRODUCTION

Myelodysplastic syndromes (MDS) are a heterogeneous group of clonal neoplasms characterized by cytopenia(s) due to ineffective hematopoiesis, dysplasia and a high-risk for AML transformation [1]. Nearly ∼50% of *de novo* MDS and ∼90% of therapy-related MDS patients are associated with characteristic structural chromosomal variants: these include (a) copy number variants (CNVs) - losses/ gains and (b) structural variants (SVs): inversions, balanced and unbalanced fusions/translocations. Identification of all structural chromosomal aberrations is crucial in clinical evaluation of MDS patients since these are essential for both diagnostic sub-classification and prognostication [1].

Firstly, per 2016 WHO classification guidelines, the diagnosis of MDS is dependent on recognition of dysplasia in at least 10% of cells in any one of the 3 lineages: granulocytic, erythroid or megakaryocytic lineages. Morphologic assessment can be challenging when the dysplasia is subtle or if the bone marrow specimen is of limited quality. On diagnostically challenging cases, detection of those MDS-defining chromosomal abnormalities proposed by the 2016 WHO criteria as “presumptive evidence of MDS” in an appropriate clinical context of unexplained cytopenia(s) is useful to establish the diagnosis of MDS [1]. Secondly, specific types of SVs can define distinct subtypes of MDS, guide treatment decisions and direct further mutation work-up, such as the use of lenalidomide therapy in MDS with isolated del(5q) in the absence of *TP53* mutations [2, 3]. Third, most importantly, structural variants represent a critical component of risk-stratification of MDS using the revised International Prognostic Scoring System (R-IPSS) [4]. The outcome of MDS is primarily dictated by the larger chromosomal aberrations detectable by karyotype despite identification of a large number of gene mutations and molecular aberrations. A comprehensive cytogenetic scoring system (CCSS), developed based on the association between chromosomal aberrations and outcome in a large cohort of MDS patients, categorizes the SVs seen in MDS into 5 different categories, ranging from “very good”, composed of del(11q) and –Y, to “very poor” encompassing >3 abnormalities. CCSS incorporates both the number and type of alterations, both copy number changes and translocations, detected by conventional karyotype alone [4, 5]. This information, together with the degree of cytopenia(s) and BM blast percentage provides the IPSS-R score for risk-stratification of MDS patients to make critical treatment decisions, both for treatment initiation and type of therapy.

Unlike somatic mutation profiling by next-generation sequencing (NGS), the development of high-throughput sequencing technologies for characterization of chromosomal scale aberrations has been limited since most of these aberrations arise in genomic regions with multiple-repeat sequences [6]. Routine clinical use of whole genome sequencing for assessment of chromosomal structural variants has not reached main stream due to the need for robust bioinformatics tools [7]. To date, for clinical work-up of MDS, most well-characterized CNVs and SVs are identified using one or more of the traditional cytogenetic techniques [4, 5, 8]. However, classical karyotyping techniques are based on evaluation of chromosomal banding patterns under light microscopy, and hence has limited resolution. Although application of high-resolution scanning of the entire genome by chromosomal microarrays for CNV analysis >30kbps has proved to be a useful diagnostic technique, it is ineffective for detection of balanced events such inversions and translocations [8-11]. Lastly, FISH techniques have limited utility due to a targeted approach.

Optical Genome Mapping (OGM) is a novel genomic technique that can potentially overcome this problem by providing a single platform for high-throughput, genome-wide detection of all the types of SVs (CNVs, balanced and unbalanced structural variants) at a high resolution [12-14]. Recent advances in OGM technology have allowed for accurate and inexpensive testing of cancer samples, including acute leukemias, for clinically relevant structural variants, all in a single assay, thereby improving turnaround times [15-18]. However, the application of OGM for clinical diagnostic work-up has seen limited adoption. Hence, as a proof-of-principle, we evaluated the performance and clinical utility of OGM-based chromosomal analysis in a series of 12 MDS patients using clinical bone marrow samples. All samples had been previously characterized by standard-of-care cytogenetic (conventional karyotype, CMA and/or FISH) and next-generation sequencing based mutation analysis. We show that OGM successfully facilitated detection and mapping of all types of structural variants with a concordance rate of 93% with conventional karyotyping and 100% with chromosomal microarray. The degree of complexity and heterogeneity in SVs characterized using OGM were not apparent using traditional techniques. Despite using stringent filtering criteria, OGM revealed 6 additional aberrations of potential clinical significance, such as *TET2* deletion, in 4 (33%) patients which were cryptic by standard-of-care technologies, majority confirmed by alternate platforms. OGM permitted precise gene-level mapping of clinically informative genes such as *TP53* and *KMT2A*, circumventing the need for multiple confirmatory assays. We conclude that OGM is a potent single-platform assay for high-throughput and accurate identification of SVs with valuable clinical utility in cancer diagnosis and identification of novel therapeutic targets.

## MATERIALS AND METHODS

### Sample Selection

We selected 12 MDS cases with available fresh/ frozen bone marrow aspirate material (5 had diploid/ normal karyotype; 7 with SVs, including 1 case showing Y chromosome loss). All cases were diagnosed as MDS or AML arising from MDS using the WHO criteria. All patients underwent conventional karyotyping, selected samples underwent chromosomal microarray analysis and FISH testing per manufacturer’s guidelines for MDS-related abnormalities in our CLIA/ CAP-certified laboratories. Clinicopathologic, cytogenetic and molecular data were reviewed. The study was approved by the Institutional Review Board and all samples were collected following institutional guidelines with informed consent in accord with the Declaration of Helsinki.

### Conventional Karyotyping and FISH studies

Conventional karyotypic studies were performed metaphase spread prepared from unstimulated 24 hour and 48 hour bone marrow aspirate cultures using standard G-banding techniques. At least 20 metaphases were evaluated, and resulted according to the 2016 International System for Human Cytogenetic Nomenclature [19, 20]. Fluorescence in situ hybridization (FISH) analysis for deletions of chromosomes 5/5q, 7/7q, +8, -17p/*TP53* and del(20q) was performed on freshly harvested aspirate smears or cultured cells with using standard techniques as previously described [21]. A total of 200 interphase nuclei were analyzed.

### Chromosomal Microarray (CMA)

CMA was done on Agilent’s custom-designed whole-genome SurePrint G3 dual-color array (4×180K chip, CCMC), with 60-mer probes [a total of 120,000 comparative genomic hybridization probes plus 60,000 single-nucleotide polymorphism probes, with ∼13 Kb genome-wide median probe spacing; the probes span >500 cancer genes and 4130 cancer-associated genomic regions]. Briefly, 500 ng of genomic DNA extracted from bone marrow aspirate samples underwent restriction enzyme digestion using *Alu* and *Rsal*, followed by Cy5-dUTP labeling using the Agilent Genomic DNA Labeling Kit Plus. For control, reference human (female) DNA (Promega Corporation, Madison, WI) was labeled with Cy3-dUTP. This was followed by hybridization per manufacturer’s recommendations. The slides were scanned using a high-resolution microarray scanner (Agilent Technologies, CA). The data analysis was done using CytoGenomics software.

### NGS-based somatic molecular profiling using the 81-gene NGS panel

All patients underwent comprehensive mutation analysis by next-generation sequencing based 81-gene panels within the CLIA-certified Molecular Diagnostics Laboratory as previously described [22]. Briefly, the genomic DNA extracted from whole mononuclear cells from fresh BM aspirates underwent amplicon-based targeted next-generation sequencing (NGS) mutation on Miseq sequencer as described previously [23, 24]. GRCh37/hg19 was used as a reference for sequence alignment. A minimum of 1% VAF with adequate coverage was required for variant calling. Since matched germline samples were not sequenced, the somatic nature of the variants was inferred based on the VAFs, evidence from the literature and online databases such as COSMIC and data from our institutional cohort. Variants reported in the Exome Aggregation Consortium [ExAC], dbSNP 137/138, and 1000 Genomes Project databases were excluded. *FLT3* ITD mutations were evaluated by PCR-based capillary electrophoresis.

### Optical Genome Mapping

Ultra-high molecular weight (UHMW) DNA was extracted from bone marrow mononuclear cells preserved in DMSO following the manufacturer’s protocols (Bionano Genomics, USA). Cells were digested with Proteinase K and RNAse A. DNA was precipitated with isopropanol and bound with nanobind magnetic disk. Bound UHMW DNA was resuspended in the elution buffer and quantified with Qubit dsDNA assay kits (ThermoFisher Scientific).

DNA labeling was performed following manufacturer’s protocols (Bionano Genomics, USA). Standard Direct Labeling Enzyme 1 (DLE-1) reactions were carried out using 750 ng of purified ultra-high molecular weight DNA. The fluorescently labeled DNA molecules were imaged sequentially across nanochannels on a Saphyr instrument. Effective genome coverage of approximately 300X was achieved for all tested samples.

Genome analysis was performed using software solutions provided by Bionano Genomics. Rare Variant Analyses were performed to sensitively capture somatic SVs occurring at low allelic fractions. Briefly molecules of a given sample dataset were first aligned against the public Genome Reference Consortium GRCh38 human assembly. SVs were identified based on discrepant alignment between sample molecules and GRCh38, with no assumption about ploidy. Consensus genome maps (*.cmaps) were then assembled from clustered sets of molecules that identify the same variant. Finally, the cmaps were realigned to GRCh38, with SV data confirmed by consensus forming final SV calls.

Finally, fractional copy number analysis was performed from alignment of molecules and labels against GRCh38 (alignmolvrefsv). A sample’s raw label coverage was normalized against relative coverage from normal human controls, segmented, and baseline CN state estimated from calculating mode of coverage of all labels. If chromosome Y molecules were present, baseline coverage in sex chromosomes was halved. With a baseline estimated, CN states of segmented genomic intervals were assessed for significant increase/decrease from the baseline. Corresponding duplication and deletion copy number variant calls were output. Certain SV and CN calls were masked, if occurring in GRC38 regions found to be high variance (gaps, segmental duplications, etc.)

### Data Analysis and Variant Filtering

Data analysis was performed in a single-blinded fashion independently by 2 users using *de novo* assembly (SVs >500bp), rare variant (SVs >10% allele fraction) and copy number (up to whole chromosome CNVs) pipelines. In order to differentiated somatic alterations from copy number variations seen in normal healthy population, we used the OGM data generated from 200 healthy controls and selected for structural abnormalities >500 bp. Based on prior sensitivity studies using simulations, serial dilutions and cell lines, a detection sensitivity of ∼95% for SVs with an allele fraction of ∼10% was achieved (Bionano Genomics, Rare Variant Pipeline).

For variant filtering, as a first step, we used the following filters to generate a list of high confidence variants for analysis. For *de novo*, rare variant and CNV pipelines, SVs were filtered based on Bionano Genomics recommended size and confidence scores (e.g. >500bp for *de novo* assembly and >5 kbp for rare variant pipelines for INDELs). CNV fractional analysis were set to be lower than <1.8 for deletions or greater than 2.2 for duplications. Additionally, CNVs below 5Mbp size cutoff were filtered out to enrich for high confidence CNV calls. In order to select clinically important aberrations, we used the following selection criteria for each of the pipelines: variants found in >1 sample within this cohort, or (2) variants that overlapped the coding region of a gene/ chromosome locus implicated in myeloid neoplasm. The list of comprehensive gene/loci was generated by combining the publicly available myeloid neoplasm-specific gene list created through a collaboration between the Cancer Genomics Consortium Education Committee and the Mayo Clinic Genomics of Oncology Annotation Team (GOAT) and our in-house 81-gene NGS mutation panel.

## Results

### Patient cohort and samples

In total, we selected BM aspirate samples from 12 MDS patients with a variety of karyotypes detected using conventional cytogenetics [4 patients with normal karyotype, 1 with a non-clonal del(9p13) in 1 of 20 metaphases (negative by chromosomal microarray analysis) and 7 patients with 1 to several chromosomal alterations]. In order to test a variety of sample types, we used 3 frozen BM cell pellets (1-4 year old pellets), 8 live BM mononuclear cells preserved in DMSO (0-1 year old) and 1 left-over BM specimen from clinical flow cytometry laboratory (at room temperature for 48 hours). The median number of cells was 1.7 (range, 0.9-5.3 x 10^6^). All samples yielded sufficient DNA (median, 54 ng/µL; range, 19-125). Irrespective of sample types, all samples with at least 1 million cells and DNA ≥36 ng/dL (as pre-determined by cell line studies), yielded successful results. The median coverage of the samples obtained was 305X.

### Concordance between OGM and standard-of-care technologies: OGM detected all types of SVs in MDS when present above the level of detection sensitivity

Our study cohort had a total of 28 clonal cytogenetic abnormalities (defined as present in 2 or more metaphases) based on clinical conventional karyotyping. These included 15 deletions, 2 insertions/ trisomies, 1 inversion, 7 translocations/ derivative chromosomes and 1 isodicentric chromosome. OGM identified 26 of 28 clonal abnormalities with a concordance rate of 89.3%. OGM was able to characterize different types of structural variants; these included copy number gains and losses in multiple chromosomes seen on the whole genome copy number profile (**Figure** 1A, sample #11), detailed mapping of the genomic coordinates of segmental deletions (**Figure** 1B circos plot showing multiple aberrations including a large 125 Mb deletion of q arm of chromosome 5 spanning from (5q11.1, 50, 250,506; 5q35.2, 174,948,964), sample #8 (**Figure** 1C, 1D) and segmental duplication of chromosome 6p (**Figure** 1E); changes in sex chromosomes (Figure 1F, loss of chromosome Y), translocations (involving chromosomes 5 and 6 in the same patient sample shown in **Figure** 1G, example of translocation involving chromosomes 2 and 22, sample #10, **Figure** 2B), inversions (**Figure** 1H, inversion involving chromosome 6; **Figure** 2A, a large ∼40Mb chromosome 3 inversion involving *MECOM/ EVI1* gene, sample #7), complex derivative chromosomes (involving segments of chromosomes 1, 12 and 5, sample #8, **Figure** 2C) and dicentric chromosome.

**Figure 1.**
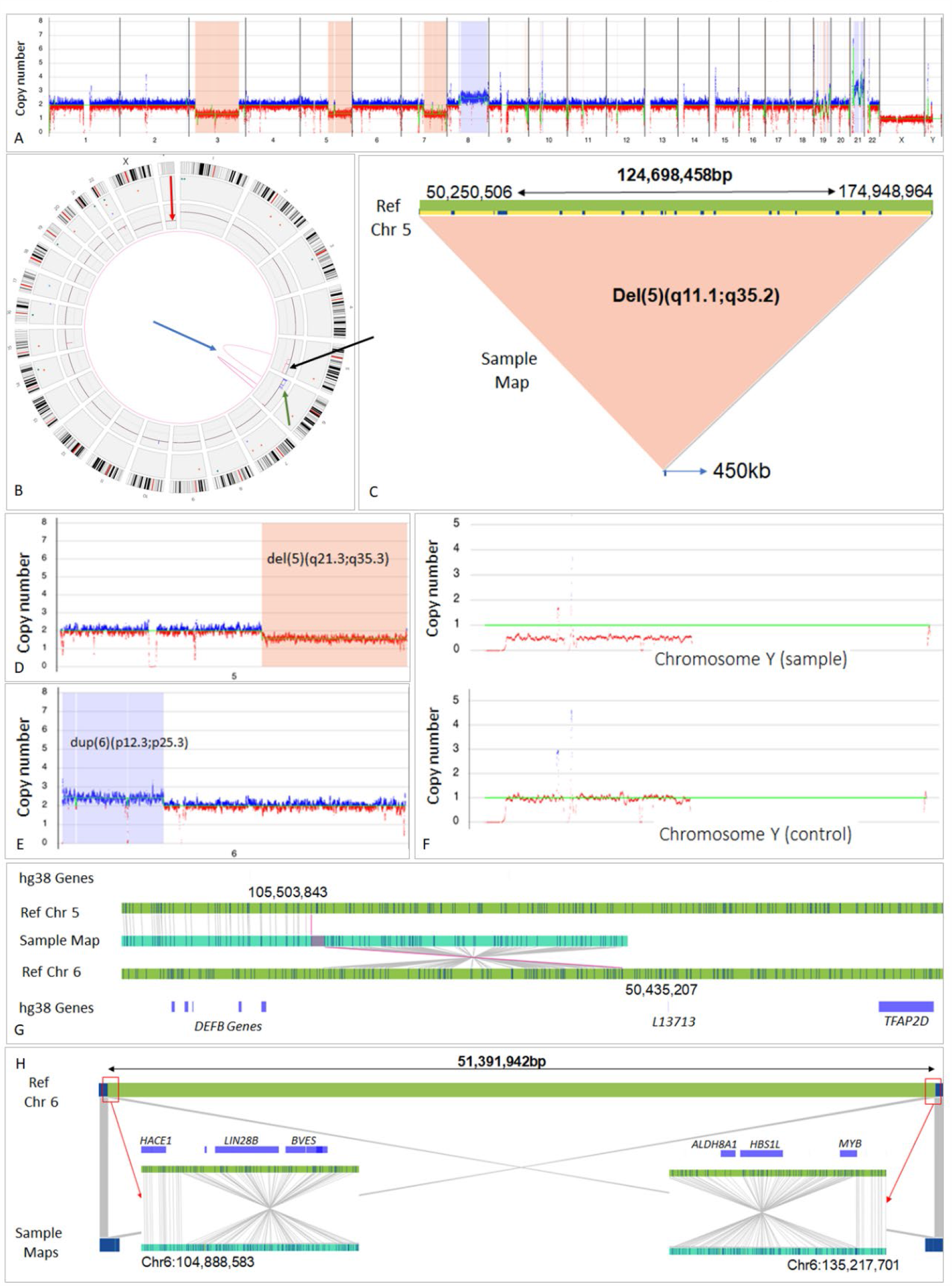
A. Whole genome copy number profiles generated by OGM. Y axis – range of copy number 0-8. X axis – human chromosomes. Molecules showing regions with increased copy number from the baseline are shown in blue and regions with decreased copy number are shown in red. This MDS sample shows chromosome 3, chromosome 5q and 7q loss, chromosome 8q gain and multiple CNVs affecting chromosome 19 and 21 (sample #11). B-H: Different types of chromosomal structural variants in MDS samples. B. Circos plot summarizing the identified genomic rearrangements and copy number profiles. The outer layer represents chromosomal G-banding locations, block underneath shows the different types of structural variants that were identified in specific locations. CNV profile is represented as the most inner block. Lines from one chromosome to another represent interchromosomal translocation events. Lines originating and ending within a single chromosome indicate intrachromosomal translocations. C. A large ∼125Mb deletion identified on the q arm of chromosome 5. Sample map in blue represents a single map spanning left and right breakpoints of the deleted region. D & E. Copy number plot showing loss of 5q (D) and gain of 6p (E) [Y axis: range of copy number 0-8; X axis: human chromosomes. Molecules showing regions with increased copy number from the baseline are in blue and regions with decreased copy number are in red]. F. Copy number plot showing copy number loss of the Y chromosome compared with a control. [Y axis: range of copy number 0-8; X axis: human chromosomes. Molecules showing regions with increased copy number from the baseline are in blue and regions with decreased copy number are in red. Top panel shows decreased Y chromosome CNV <1. Bottom panel shows an unrelated control male with Y chromosome copy number at 1. Chromosome Y loss was not readily apparent in the circos plot]. G & H. Translocation (G) and inversion (H). GRCh38 reference chromosomes with OM label patterns are shown in green. Assembled sample maps with label patters are shown in light blue. Label alignments between two maps are shown in grey strings. Translocation breakpoints are highlighted in purple. Overlapping genes are shown in blue.

**Figure 2.**
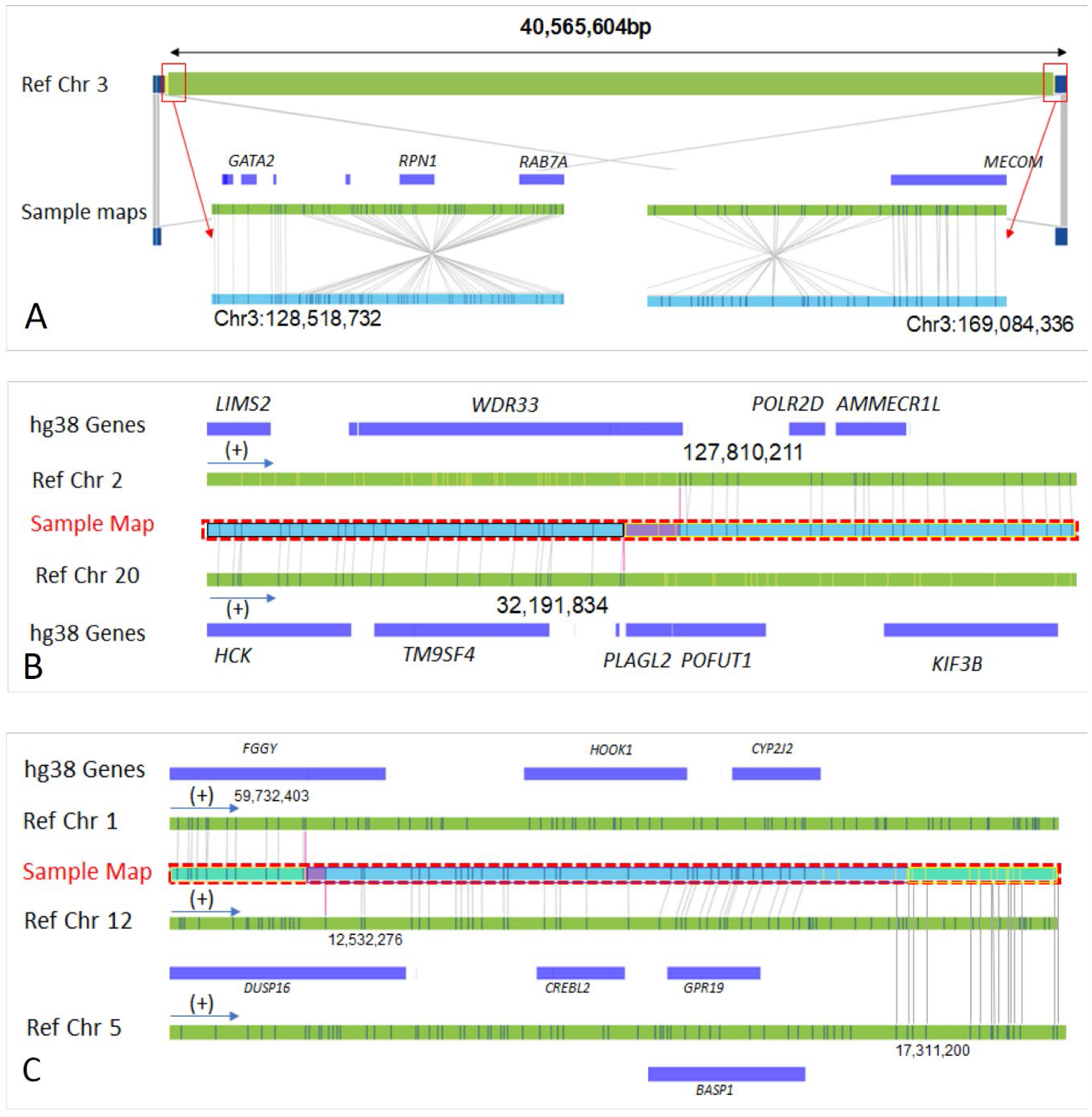
**A**. Large ∼40Mb chromosome 3 inversion identified in MDS, sample #7. GRCh38 reference chromosomes with OM label patterns are shown in green. Assembled sample maps with label patters are shown in light blue. Label alignments between two maps are shown in grey strings. Overlapping genes are shown in blue. B. A translocation event involving chromosomes 2 and 22 identified in MDS, sample #10. GRCh38 reference chromosomes with OM label patterns are shown in green. Assembled sample maps with label patters are shown in light blue. Label alignments between two maps are shown in grey strings. Translocation breakpoints are highlighted in purple. Overlapping genes are shown in blue. C. A complex translocation event (derivative chromosome) involving chromosomes 1, 12 and 5 identified in MDS, sample #8. GRCh38 reference chromosomes with OM label patterns are shown in green. Assembled sample maps with label patters are shown in light blue. Label alignments between two maps are shown in grey strings. Translocation breakpoints are highlighted in purple. Overlapping genes are shown in blue.

The 2 chromosomal aberrations missed by OGM included add(5)(q35) and add(22)(q13)/+22, both were seen in a single patient within a composite karyotype of 4 cells. Hence, both these aberrations were likely present at a level below the level of detection. Upon reviewing the raw molecule data, segmental CNVs of small size were observed in chromosomes 22 (22q13, 10 kb insertion) and 5 (30 kb deletion), but were not present at sufficient confidence levels and did not meet our filtering cut-off. Dic(9;18)(p13;p11.2) seen in all 20 metaphases, although initially missed using standard filtering criteria since the breakpoints overlapped with genomic regions of segmental duplication, but was apparent on viewing raw molecules. Hence, particular attention to filtering criteria was needed to regions showing segmental duplication.

Overall, OGM serves as a single platform assay that can identify different types of structural chromosomal alterations of potential clinical significance detected using karyotype and CMA. We have summarized the findings of OGM compared to other clinical diagnostic techniques in **Table 1**. Representative images of each type of aberration illustrated in **Figures** 1 and 2.

**Table 1.**
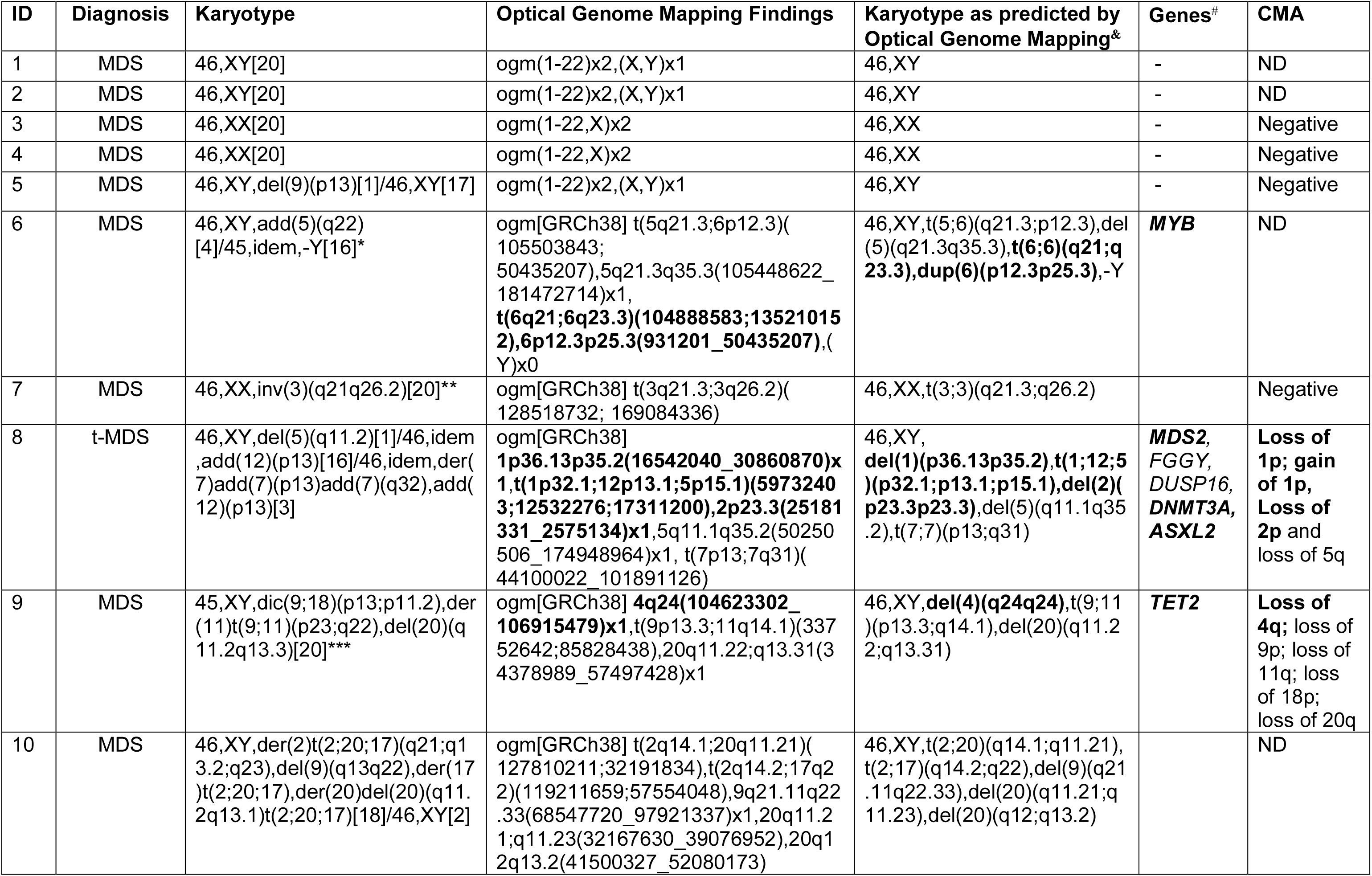
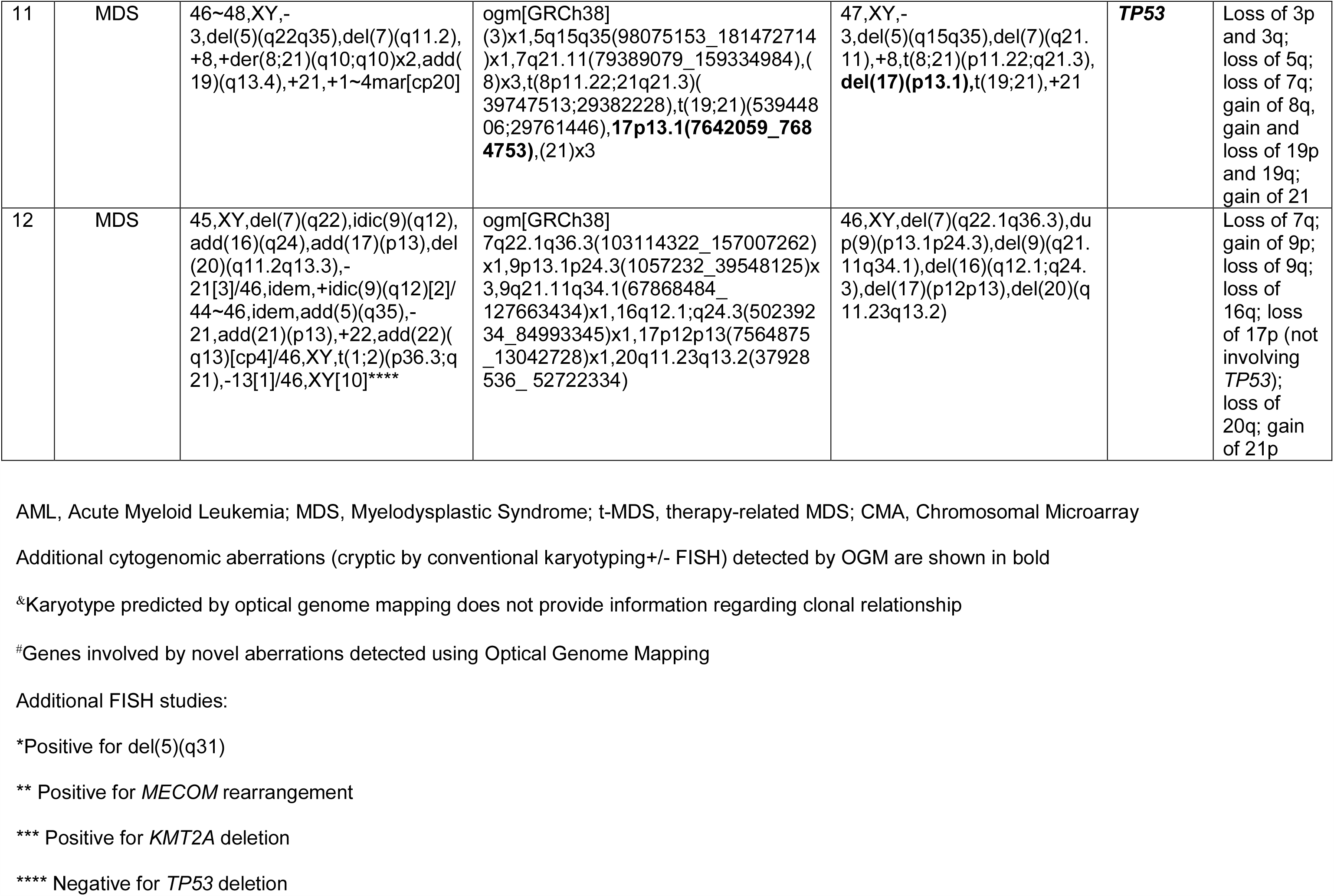
Summary of all chromosomal aberrations seen in the MDS study cohort at baseline using optical genome mapping (OGM) compared to conventional karyotyping and/or FISH and chromosomal microarray. In addition to concordance, cryptic aberrations uncovered by OGM is in bold.

### OGM revealed additional clinically relevant sub-microscopic aberrations that were cryptic by standard-of-care technologies

Aside from confirming the cytogenetic aberrations detected using conventional techniques, OGM identified several additional chromosomal alterations. These are listed in **Table 1**. For the purpose of this study, we focused on structural variants that were of potential clinical importance. In order to filter for variants with highest likelihood of clinical significance, we applied stringent criteria that included the following: (1) variants noted in >1 sample within this cohort, or variants that overlapped the coding region of a gene/ chromosome locus implicated in myeloid neoplasm. The list of comprehensive gene/loci was generated by combining the publicly available myeloid neoplasm-specific gene list created through a collaboration between the Cancer Genomics Consortium Education Committee and the Mayo Clinic Genomics of Oncology Annotation Team (GOAT) and our in-house 81-gene NGS mutation panel [24]. After excluding the aberrations restricted to intronic regions, we identified 6 potentially significant aberrations in 4 (33%) patients that were cryptic by conventional karyotype (see **Table 1**). These included deletions of chr 1 (*MDS2*), chr 2 (*ASXL2, DNMT3A*), chr 4 (*TET2*), chr 17 (*TP53*), duplication involving chr 1 (*RPL22, CSMD2, CSF3R, JAK1* and *GFI1*) and translocation involving chr 6 (*MYB*). None of the diploid karyotype MDS showed additional abnormalities. The significance of each of these abnormalities are described in subsequent sections in appropriate clinical contexts.

### High-resolution OGM uncovered the complexity of structural alterations and heterogeneity in breakpoint regions that were not apparent using conventional techniques

For known cytogenetic aberrations detected using standard techniques, OGM was able to provide more precise mapping of genomic coordinates coupled with gene level information, which is critical to evaluate the clinical relevance. OGM uncovered the complexity of the structural variants that was not apparent by conventional karyotype. To illustrate, we show the heterogeneity in SVs using 3 different MDS cases, all of which showed deletions of chromosome 5 (cases #6, #8 and #11) by conventional karyotype. Both cases #11 and #6 involved deletions of critically important genes: Case #11 was a complex karyotype MDS with segmental deletion of chr5. In addition to confirming these aberrations, OGM identified a *TP53* deletion not picked up by karyotype. Case #6, was a low-grade *SF3B1* mutated MDS progressing to AML, with segmental deletion of chr5 and loss of Y by karyotype. By OGM, it was evident that the segmental chr 5 deletion (involving several genes including *EGR1, PDGFRB, RPS14, NPM1, NSD1, DDX41*) was associated with an additional translocation t(5;6)(q21.3;p12.3) involving *MYB* gene and duplication of chr6 involving *JARID2* and *DEK* genes. The latter two aberrations were not detectable by karyotype. Genomic gain in 6p21 has been shown to be major pathogenic events in progression of MDS to AML [25]. Case #8 was a therapy-related complex karyotype MDS, with segmental deletions of chromosomes 5, 7 and 12. In addition to confirming these, OGM mapped a complex structural aberration composed of fragments of chromosomes 1, 5 and 12 leading to a complex derivative chromosome t(1;12;5)(p32.1;p13.1;p15.1) that could not be discerned by any of the conventional techniques, along with cryptic deletion (*MDS2*) and duplication of chr 1. Aside from this, heterogeneity of breakpoint regions within the same chromosome 20 shown by multiple molecules representing different cell populations was demonstrated by OGM (case #9).

### Precise mapping at the gene-level in OGM is clinically relevant and informative for clinical management of MDS and voids the need for multiple confirmatory assays

The inherent higher resolution of OGM with more precise mapping of genomic coordinates is helpful to resolve gene-level details in the aberrations that are pertinent for clinical management. Recent study by Bernard et al. has shown that multi-hit alterations of *TP53* gene (defined as multiple *TP53* mutations, or *TP53* mutation with either *TP53* deletion or CN-LOH, all indicative of multiallelic alterations) in MDS are biologically and clinically distinct from monoallelic *TP53* alterations (defines as a single *TP53* mutation alone) [26]. Determination of *TP53* allelic state is clinically important. Only MDS patients with multi-hit *TP53* state are associated with poor outcome and treatment resistance, especially to hypomethylating agents while the prognosis and therapy responses of MDS patients with *TP53* monoallelic state patients are similar to those with wild-type *TP53* [26]. Currently, mutation data is evaluated by routine targeted NGS sequencing. The second hit of *TP53* can be either deletion, CN-LOH or a rearrangement, this data is usually obtained by karyotype and/or CMA; however, latter cannot identify rearrangements. Confirmation of *TP53* gene deletion from karyotype needs additional FISH studies, since the chromosomal morphology is often limited by the complexity in the karyotype. OGM can provide the gene-level information to evaluate the status of the second *TP53* hit by any type of alteration, upfront, without the need for additional confirmation studies, as illustrated in 2 examples below.

Case #11 was a *TP53* mutated (p.M237I, 72% VAF) MDS patient with a complex karyotype showing no chromosome 17 alterations. Chromosomal microarray testing by two different platforms confirmed multiple alterations on chromosomes 3, 5, 7 among others but no alteration in chromosome 17. OGM uncovered a cryptic 1.5 kb deletion of chromosome 17 involving the *TP53* gene. Based on this information, *TP53* allelic state would be regarded as “multi-hit”. On the other hand, case #12 was a *TP53* mutated MDS patient (R248W, 55% VAF) with complex karyotype showing additional material on chromosome 17(p13) [add(17)(p13)] in at least 7 of 20 metaphases, suggestive of a concurrent *TP53* deletion. However, all 3 assays (FISH, chromosomal microarray and OGM) confirmed the absence of *TP53* deletion. OGM further clarified that there was a 7.5 Mb deletion in the p-arm of chromosome 17 spanning multiple genes including *PRPF8* and *RABEP1*, located proximal to *TP53* gene but not involving it, making this a mono-allelic *TP53* hit. In addition to *TP53*, OGM also allowed determination of clinically important findings in other genes, such as *KMT2A* and *TET2*, as described in the next section.

### OGM, as a single-platform assay for accurate identification of all types of structural variants underscored by a case example

Finally, we illustrate the biological and clinical implications of identification of cryptic structural aberrations at a high resolution in a comprehensive manner using a single-platform assay using an example of an MDS patient, with conventional karyotype at the time of diagnosis showing a derivative chromosome 11 among other abnormalities, including t(9;11): 45,XY,dic(9;18)(p13;p11.2),der(11)t(9;11)(p23;q22),del(20)(q11.2q13.3)[20]. The presence of t(9;11) suggested the involvement of *KMT2A/MLL* gene. While OGM confirmed the presence of t(9;11)(p13.3;q14.1), the fusion, however, did not involve the *KMT2A* gene, but was associated with a 49.3 Mbp segmental deletion of chromosome 11 encompassing *KMT2A, PICALM, EED, CBL* and multiple other genes. FISH studies using a dual color breakapart rearrangement probe (Abott Molecular, Inc) confirmed the deletion of *KMT2A* gene in 91% cells and the absence of *KMT2A* rearrangement. This result was also confirmed using map-back FISH studies on the previously G-banded karyotyped metaphases. *KMT2A* rearrangements (which are characteristic of therapy-related myeloid neoplasms associated with exposure to prior topoisomerase 2 inhibitor therapy) and *KMT2A* deletions have distinct clinicopathologic characteristics and prognostic implications in MDS [1, 27]. The more important point to note is that the precise gene-level information regarding fusions as well as deletions was obtainable by OGM assay alone, and may abrogate the need for confirmatory FISH studies. In this patient, OGM further uncovered a cryptic t(4;7) fusion associated with a cryptic 2.3 Mbp segmental deletion of chromosome 4 involving *TET2* gene. *TET2* deletions are frequent in MDS but often cryptic by karyotype. Putting hemizygous *TET2* deletion together with the NGS studies that showed *SF3B1* K700E mutation and a frame-shift *TET2* mutation (pS128fs), the findings demonstrate a biallelic *TET2* inactivation. Biallelic *TET2* inactivation is common in lower-risk MDS with monocytosis, but not meeting the current WHO criteria for chronic myelomonocytic leukemia (CMML), and a low-risk for leukemia transformation [28]. All of the above findings bear significance in MDS prognostication and treatment decisions. From a laboratory perspective, OGM, as a single-platform assay, voided the need for confirmatory FISH testing for *KMT2A* and *TET2* in these scenarios (**Figure** 3, A-E). These findings show that OGM is a potent technology for gathering these additional layers of data regarding genomic complexity for further studies on clinical correlation.

**Figure 3.**
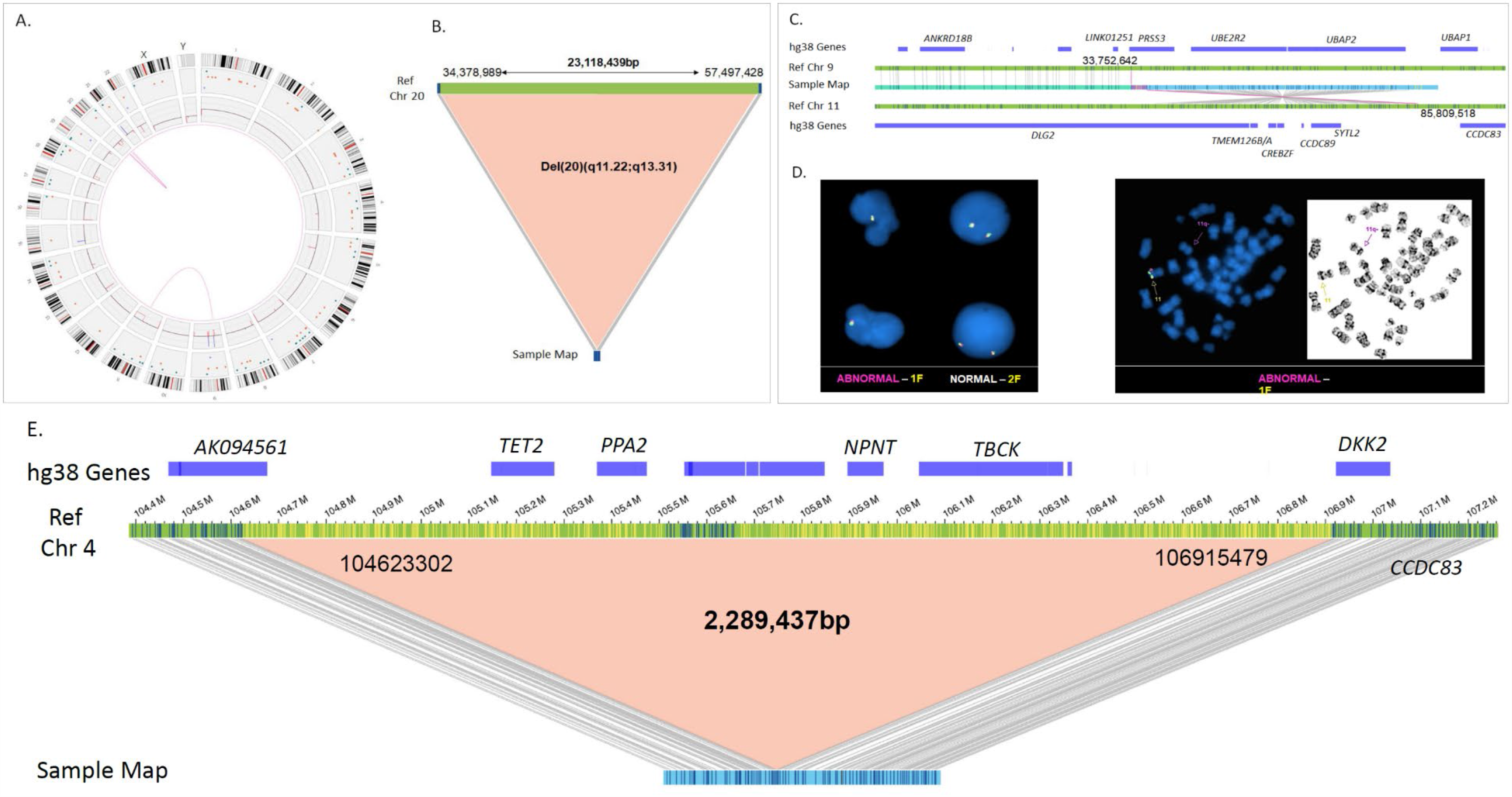
MDS Case # 9. A. Circos plot summarizing the identified genomic rearrangements and copy number profiles. B. Large ∼23Mb deletion identified on the q arm of chromosome 20. C. A translocation event involving chromosomes 9 and 11, but not involving *KMT2A* gene. D. There was a concurrent 49 Mbp deletion encompassing the *KMT2A* gene, FISH studies using LSI KMT2A/MLL dual color breakapart rearrangement probe (Abott Molecular, Inc) and map-back FISH done on previously G-banded and karyotyped metaphases confirmed the absence of *KMT2A* fusion and deletion of 1 copy of the gene. E. In addition, approximately 2Mb cryptic deletion was identified by OGM on chromosome 4 that includes *TET2* gene.

## DISCUSSION

In the last decade, next-generation-sequencing for gene mutation analysis has defined the complexity in the genomic landscape of cancer. Congruent advances in detection of structural variants that are of equal significance, if not more, was lacking due to the presence of long replicating complex genomic regions. OGM largely overcomes these problems due to the ability to map long DNA molecules, and represents a significant tool in advancing cytogenomic profiling of cancer.

For this proof-of-principle study, we selected MDS as a prototype for hematological malignancies for the following reasons: (1) nearly ∼50% of *de novo* MDS and ∼90% of therapy-related MDS patients are associated with a structural variant. MDS is associated with characteristic chromosomal abnormalities (2) detection of SVs is essential for risk stratification including R-IPSS scoring, and guiding treatment decisions, as well as for establishing the diagnosis in difficult clinical scenarios based on MDS-defining “presumptive” cytogenetic abnormalities (3) Further, MDS is a precursor to a significant proportion of AML with cytogenetic aberrations; hence, this provides an opportunity to understand the karyotypic evolution at a high-resolution.

Based on our preliminary analysis, we have verified the feasibility and robust performance of OGM on a variety of bone marrow samples we encounter both for clinical and research purpose. The feasibility of OGM testing, especially on the left-over (to be discarded) BM specimen in our flow cytometry laboratory is particularly helpful. At the onset, testing may be reserved for diagnostically challenging cases or cases that are refractory to therapy, until the cost per sample for OGM becomes comparable to conventional karyotype.

Since OGM can generate a large number of variants, we have adopted strict filtering criteria to determine those SVs that overlapped regions encompassing genes known to be implicated in MDS pathogenesis. Based on this, as evident from our study, OGM uncovered 6 additional cryptic clinically important chromosomal alterations involving prognostic genes such as *TP53* and *KMT2A*, all of which were subsequently confirmed using FISH and/or CMA. The clinical significance of each of these have been already described under the respective sections [26-29]. These preliminary findings need further validation on a larger scale in uniformly treated MDS patients. Further, relaxing the filtering criteria would enable discovery of additional genomic in understanding the disease pathology and progression, beyond the mutations.

OGM has a few limitations. There is a need to develop algorithms for detection of detect copy-neutral loss of heterozygosity. Interpretation of OGM results is technically less laborious, and the need for confirmatory tests is largely mitigated once validation has been completed. Both of these factors partly offset the higher cost-per-sample compared to karyotype at this time. Since OGM requires long molecules (150 kbp) to aid assembly of long replicating complex regions, gentle processing, handling and storage of samples is required. Nevertheless, the broad applications of OGM testing fits with the clinical laboratory workflow setting.

In summary, OGM is a powerful and reliable single-platform cytogenomics tool for high-throughput detection of all types of clinically important SVs in myeloid malignancies.

## Data Availability

All data have been reported in the attached tables and figures.

